# The association between nurse staffing configurations and sickness absence: longitudinal study

**DOI:** 10.1101/2024.09.02.24312931

**Authors:** Chiara Dall’ora, Paul Meredith, Christina Saville, Jeremy Jones, Peter Griffiths

**Author notes:** Corresponding author: Dr Chiara Dall’Ora; School of Health Sciences, University of Southampton; Room 3033, Building 67, Highfield Campus; Southampton SO17 1BJ.

## Abstract

**Importance:** Nurses’ work-related stress and sickness absence are high. The consequences of sickness absence are severe for health systems’ efficiency and productivity.

**Objective:** To measure the association between nurse staffing configurations and sickness absence in hospital ward nursing teams.

**Design:** Retrospective case-control study using hospital routinely collected data

**Setting:** Four general acute care hospitals in England

**Participants:** 3,583,586 shifts worked or missed due to sickness absence by 18,674 registered nurses (RN) and nursing assistant (NA) staff working in 116 hospital units.

**Exposure:** Nursing team skill-mix; temporary staffing hours; understaffing; proportion of long shifts (12+ hours) worked; full-time/part-time work status in the previous 7 days.

**Main outcome:** Episodes of sickness absence, defined as a sequence of sickness days with no intervening days of work.

**Results:** There were 43,097 sickness episodes. In our reduced parsimonious model, being exposed to a skill mix that was richer in RNs was associated with lower RN sickness absence (OR= 0.98; 95% CI = 0.96-0.99). For each 10% increase in proportion of hours worked as long shifts worked in the previous 7 days odds of sickness were increased by 2% (OR = 1.02; 95% CI = 1.02- 1.03) for RNs. Part-time work for RNs was associated with higher sickness absence (OR = 1.09; 95% CI = 1.04 – 1. 15). When RN staffing over the previous week was below average, the odds of sickness absence for NAs increased by 2% for every 10% increase in understaffing across the period (OR = 1.02; 95% CI = 1.01 - 1.03). For RNs there was a significant interaction between part-time work and RN understaffing, whereby short staffing in the previous week increased sickness absence for full time staff but not among those working part time. NA understaffing was not associated with sickness absence for any staffing group.

**Conclusions and Relevance:** Working long shifts and working on understaffed wards increases the risk of sickness absence in nursing teams. Adverse working conditions for nurses, already known to pose a risk to patient safety, may also create risks for nurses and the possibility of further exacerbating staff shortages.

**Key points:** *Question:* What is the association between variation in nurse staffing configurations and nurses’ sickness absence?

*Findings:* Registered Nurse (RN) understaffing in the preceding 7 days was associated with sickness absence for Nursing Support (NS) staff, but for RNs the association was only seen when working full time. Exposure to shifts with a skill-mix richer in RNs, to higher bank hours and working lower proportions of 12+ h shifts in the preceding 7 days was a protective factor of RN sickness absence.

*Meaning:* To support nurses’ health and health systems’ productivity and efficiency, investing in avoiding RN understaffing may be warranted.

## Introduction

The nursing workforce worldwide is experiencing high levels of work-related stress and sickness absence, with concerns around nurses’ ability to cope with such high pressure in the long term.^1^ Every year, England’s NHS nursing staff report considerable dissatisfaction with their jobs and feelings of burnout,^2^ with sickness absence levels remaining higher than those of most other health professions.^3^

The implications of high sickness absence levels are serious, both in terms of lost productivity and increased costs to employers and the society.^4^ Mental health-related complaints, including stress and anxiety, are the main cause of sickness absence in nursing staff in England, making up around a quarter of all absences.^3^ While these figures do not offer insights as to the causes of such mental health-related complaints, in the NHS staff survey nearly half the nurses reported they felt unwell because of work-related stress in the last 12 months.^2^

While sickness absence is a complex phenomenon, not always caused by occupational factors, there is evidence that several modifiable work-related factors are associated with nurses’ sickness absence, in particular working long shifts.^5,6^ Nurse staffing levels have been associated with a range of staff outcomes in cross-sectional survey studies, including burnout, job dissatisfaction, intention to leave one’s job and the profession, ^7,8^ but the impact on sickness absence rates has not been studied directly. Similarly, a lower registered nurse (RN) skill mix (i.e. lower proportions of RNs within the nursing team configuration) and working on shifts with higher levels of temporary bank and agency staff have been associated with increased turnover^9^ and job dissatisfaction^10^ but the impact on sickness absence has not been studied.

This gap is likely to have originated in part from unavailability of objective sickness absence data, but advances in research using data extracted from routinely collected electronic hospital systems mean that this gap can now be addressed. Therefore, the aim of this study was to measure the association between nurse staffing configurations and nurse sickness absence in general acute care hospitals.

## Methods

This was a retrospective longitudinal study in four NHS hospital Trusts across England. These Trusts display variation in terms of being geographically located across England (i.e., London, Southeast, Southwest, Midlands), in areas with mixed urban and rural catchments (i.e., three hospitals are urban, one is rural), and different levels of affluence and deprivation. They also exhibit variation in Trust type (two are teaching hospitals), size (ranging from 523 to 1746 beds), average RN staffing levels (ranging from 1.38 to 2.74 RNs per occupied bed), making the sample heterogeneous. We included all adult acute inpatient wards / admissions units (including High Dependency Units and Intensive Care Units).

Our data sources were the electronic rostering systems, containing records of all nursing staff rostered to work on a ward on a given shift and records of temporary bank (i.e., hospital-employed staff working extra hours beyond their contract) and external agency staff working on the ward, and the hospitals’ patient administration systems. We used the latter to quantify how many patients were on a ward at any given point in time. Data were available from April 2015 to February 2020 in most Trusts, except for one Trust where data were available from March 2019. In total, we extracted 3,583,586 shifts from 123 wards. Staff identifiers in rostering records were pseudonymised allowing the linkage of shifts worked by the same individual across the study period and linkage to sickness absences. Sickness absence data were available for employed staff only, not for bank or agency staff. Shifts recorded as sickness absence were aggregated to identify sickness episodes. A sickness episode started on the first day a nurse was absent from work and finished as soon as they returned to work for at least one shift. We removed episodes that were not preceded by any worked hours in the previous 7 days.

We calculated ward staffing levels for 12-hour shift periods starting at 7am (day) and 7pm (night) by aggregating rostered nursing hours of care, including temporary assignments to the ward (i.e., bank and agency). We calculated staffing hours excluding breaks by assuming breaks were taken in the middle of shifts. We then linked the nursing hours to the patient occupancy on the ward for the same 12-hour periods. This was achieved by calculating the occupancy for each patient on the ward during the shift using the admission, discharge, and transfer information for the ward, aggregating the occupancy durations, and then converting the total occupancy duration into a number of patient-days. We calculated hours per patient day (HPPD) for each shift as the sum of hours worked by each staff group divided by patient-days. We calculated RN and nursing support (NS) staffing levels separately using the NHS Agenda for Change pay bands to identify RNs (band 5+) and NSs (bands 2 – 4). NSs work under the guidance of RNs and support them in the delivery of nursing services. We defined an expected staffing level by shift period (day or night) for each staff group on each ward by calculating the mean staffing, with the expected staffing rebased when there were clear discontinuities in ward use, identified by substantial changes in case-mix. Rebasing involved calculating ward staffing means for each consistent period of case-mix around a discontinuity. Shifts with unfeasibly low (fewer than 11 hours of RN staffing in a 12-hour period or less than 0.5 RN HPPD) or high (more than 48 RN HPPD) staffing were excluded. These were assumed to arise due to a mismatch between patient and staff records occurring during periods of ward reorganisation.

For each staff member for each shift we calculated various exposure measures to staffing configurations for all shifts they had worked in the past seven days. These exposure measures were understaffing of RN and NS staff, the proportion of staff hours provided by temporary bank and agency staff on the ward, the skill mix (proportion of all hours provided by RNs) and the proportion of long shifts (≥ 12-hours) worked by that individual. Understaffing was calculated as the mean of 1- (observed HPPD / expected HPPD) where observed HPPD was smaller than expected i.e., as a unidirectional measure. We classified an employee as part-time when their median weekly worked hours in the previous 13 weeks was less than or equal to 26 hours.

We explored the association between staffing configurations and sickness absence with generalized linear mixed models with a logit link function. We calculated intraclass correlation coefficients (ICC) from unconditional random intercept models to assess the within-ward and the within-staff variation for sickness. There was variation in sickness episodes at the individual level (ICC = 0.85) and at the ward level (ICC = 0.49), so both were included as random effects. All analyses were performed at the shift level. Because sickness absence rates differ substantially for registered nurses and nursing assistants, we modelled them separately.^3^ To aid interpretations of results, we input understaffing, bank and agency, skill mix and long shift variables as 10 percentage point increments in our models. We estimated univariable models (single staffing factor), a full model (all staffing factors) and a parsimonious reduced model, using backwards stepwise selection, removing the variables with the highest p-value at each step, provided this would lead to reductions in AIC and BIC. In the backwards stepwise selection, we did not remove RN understaffing because it was a theoretical focal point of our analysis. Because the effect of RN understaffing on sickness absence might depend on other variables, including NS understaffing, bank and agency hours and skill mix, we tested for interactions between RN understaffing and the above variables. To exclude multicollinearity, we checked the variance inflation factor (VIF) of all models; VIF scores were <10, indicating low multicollinearity.^11^ Data analyses were undertaken using R,^12^ and the lme4 package.^13^ This study received ethical approval by the Health Research Authority (IRAS 273185) and the University of Southampton ethics committee (ERGO 52957).

## Results

After removing all shifts with unrealistic staffing levels, our sample was 2,690,080 shifts in 116 wards, of which 43,097 were the first day of a sickness episode. In total there were 18,674 members of staff. 2,188,562 (81.6%) were shifts by staff classified as working full time, and 493,400 (18.4%) by staff classified as working part-time.

Descriptive statistics for exposure to staffing configurations in the past week by sickness cohort are reported in Table 1.

**Table 1.** Staffing configurations by sickness cohort.

| Exposure in the past week |  | All groups |  | RNs |  | NSs |  |
| --- | --- | --- | --- | --- | --- | --- | --- |
|  |  | Sick | Worked | Sick | Worked | Sick | Worked |
| **RN understaffing** |  |  |  |  |  |  |  |
|  | Median | 6% | 5% | 4.8% | 5% | 10% | 10% |
|  | Mean | 9% | 9% | 8.2% | 8% | 15% | 15% |
| **NA understaffing** |  |  |  |  |  |  |  |
|  | Median | 8% | 8% | 6.8% | 7% | 5.7% | 6% |
|  | Mean | 13% | 14% | 10.4% | 10% | 10.9% | 11% |
| **Agency staffing (proportion)** |  |  |  |  |  |  |  |
|  | Median | 0% | 0% | 0% | 0% | 0% | 0% |
|  | Mean | 4% | 3% | 3% | 2% | 4% | 3% |
| **Bank staffing (proportion)** |  |  |  |  |  |  |  |
|  | Median | 0% | 0% | 0% | 0% | 0% | 0% |
|  | Mean | 7% | 7% | 7.1% | 6% | 7.8% | 8% |
| **Skill mix (proportion)** |  |  |  |  |  |  |  |
|  | Median | 61% | 63% | 66% | 68% | 55% | 56% |
|  | Mean | 63% | 64% | 67% | 68% | 56% | 56% |
| **Long (≥12-h) shifts (proportion)** |  |  |  |  |  |  |  |
|  | Median | 100% | 100% | 100% | 100% | 100% | 100% |
|  | Mean | 79% | 78% | 81.2% | 78.2% | 76.5% | 78% |

All odds ratios and confidence intervals of associations between staffing configurations and sickness absence are reported in **Table 2**.

**Table 2.**
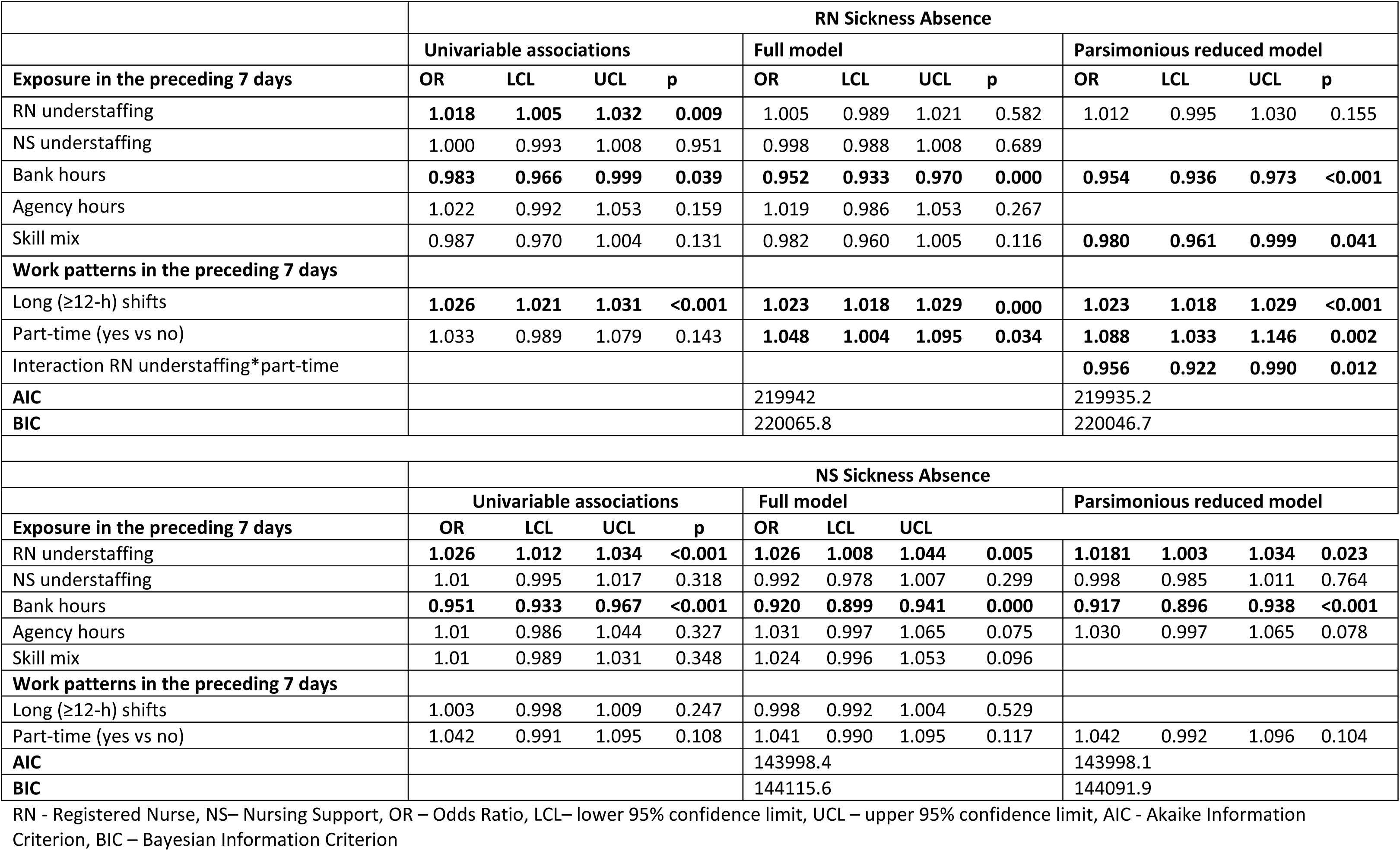
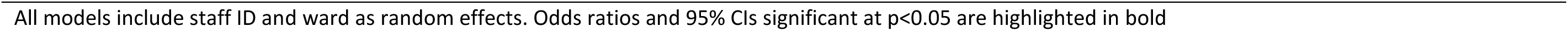
Regression – univariable and multi variable associations between staffing configurations and sickness absence.

|  | RN Sickness Absence |  |  |  |  |  |  |  |  |  |  |  |
| --- | --- | --- | --- | --- | --- | --- | --- | --- | --- | --- | --- | --- |
|  | Univariable associations |  |  |  | Full model |  |  |  | Parsimonious reduced model |  |  |  |
| Exposure in the preceding 7 days | OR | LCL | UCL | p | OR | LCL | UCL | p | OR | LCL | UCL | p |
| RN understaffing | **1.018** | **1.005** | **1.032** | **0.009** | 1.005 | 0.989 | 1.021 | 0.582 | 1.012 | 0.995 | 1.030 | 0.155 |
| NS understaffing | 1.000 | 0.993 | 1.008 | 0.951 | 0.998 | 0.988 | 1.008 | 0.689 |  |  |  |  |
| Bank hours | **0.983** | **0.966** | **0.999** | **0.039** | **0.952** | **0.933** | **0.970** | **0.000** | **0.954** | **0.936** | **0.973** | **<0.001** |
| Agency hours | 1.022 | 0.992 | 1.053 | 0.159 | 1.019 | 0.986 | 1.053 | 0.267 |  |  |  |  |
| Skill mix | 0.987 | 0.970 | 1.004 | 0.131 | 0.982 | 0.960 | 1.005 | 0.116 | **0.980** | **0.961** | **0.999** | **0.041** |
| Work patterns in the preceding 7 days |  |  |  |  |  |  |  |  |  |  |  |  |
| Long (≥12-h) shifts | **1.026** | **1.021** | **1.031** | **<0.001** | **1.023** | **1.018** | **1.029** | **0.000** | **1.023** | **1.018** | **1.029** | **<0.001** |
| Part-time (yes vs no) | 1.033 | 0.989 | 1.079 | 0.143 | **1.048** | **1.004** | **1.095** | **0.034** | **1.088** | **1.033** | **1.146** | **0.002** |
| Interaction RN understaffing*part-time |  |  |  |  |  |  |  |  | **0.956** | **0.922** | **0.990** | **0.012** |
| AIC |  |  |  |  | 219942 |  |  |  | 219935.2 |  |  |  |
| BIC |  |  |  |  | 220065.8 |  |  |  | 220046.7 |  |  |  |
|  | NS Sickness Absence |  |  |  |  |  |  |  |  |  |  |  |
|  | Univariable associations |  |  |  | Full model |  |  |  | Parsimonious reduced model |  |  |  |
| Exposure in the preceding 7 days | OR | LCL | UCL | p | OR | LCL | UCL |  |  |  |  |  |
| RN understaffing | **1.026** | **1.012** | **1.034** | **<0.001** | **1.026** | **1.008** | **1.044** | **0.005** | **1.0181** | **1.003** | **1.034** | **0.023** |
| NS understaffing | 1.01 | 0.995 | 1.017 | 0.318 | 0.992 | 0.978 | 1.007 | 0.299 | 0.998 | 0.985 | 1.011 | 0.764 |
| Bank hours | **0.951** | **0.933** | **0.967** | **<0.001** | **0.920** | **0.899** | **0.941** | **0.000** | **0.917** | **0.896** | **0.938** | **<0.001** |
| Agency hours | 1.01 | 0.986 | 1.044 | 0.327 | 1.031 | 0.997 | 1.065 | 0.075 | 1.030 | 0.997 | 1.065 | 0.078 |
| Skill mix | 1.01 | 0.989 | 1.031 | 0.348 | 1.024 | 0.996 | 1.053 | 0.096 |  |  |  |  |
| Work patterns in the preceding 7 days |  |  |  |  |  |  |  |  |  |  |  |  |
| Long (≥12-h) shifts | 1.003 | 0.998 | 1.009 | 0.247 | 0.998 | 0.992 | 1.004 | 0.529 |  |  |  |  |
| Part-time (yes vs no) | 1.042 | 0.991 | 1.095 | 0.108 | 1.041 | 0.990 | 1.095 | 0.117 | 1.042 | 0.992 | 1.096 | 0.104 |
| AIC |  |  |  |  | 143998.4 |  |  |  | 143998.1 |  |  |  |
| BIC |  |  |  |  | 144115.6 |  |  |  | 144091.9 |  |  |  |

RN - Registered Nurse, NS– Nursing Support, OR – Odds Ratio, LCL– lower 95% confidence limit, UCL – upper 95% confidence limit, AIC - Akaike Information Criterion, BIC – Bayesian Information Criterion
All models include staff ID and ward as random effects. Odds ratios and 95% CIs significant at $p < 0.05$ are highlighted in bold

In our reduced parsimonious model, considering RN sickness absence, for each additional 10 percentage points of hours worked as long shifts by an RN in the prior 7 days, there was a 2% increase in the odds of sickness absence (OR= 1.02; 95% CI = 1.02-1.03). For every 10 percentage points increase in exposure to bank hours on the ward in the previous week, there was a 5% reduction in the odds of sickness absence (OR = 0.95; 95% CI = 0.936 - 0.973). Skill mixes that were richer in RNs were associated with lower sickness absence (for every 10 percentage points increase in proportions of RNs in the previous week: OR = 0.98; 95% CI = 0.96 – 0.99). Working part-time was associated with higher sickness absence (OR= 1.088; 95% CI = 1.033 - 1.146). We found a statistically significant interaction (p=0.012) between exposure to RN understaffing and working part-time. To understand the interaction, we plotted the curves based on the B coefficients of RN understaffing, part-time work and of the interaction (see **Figure 1**).

**Figure 1.**
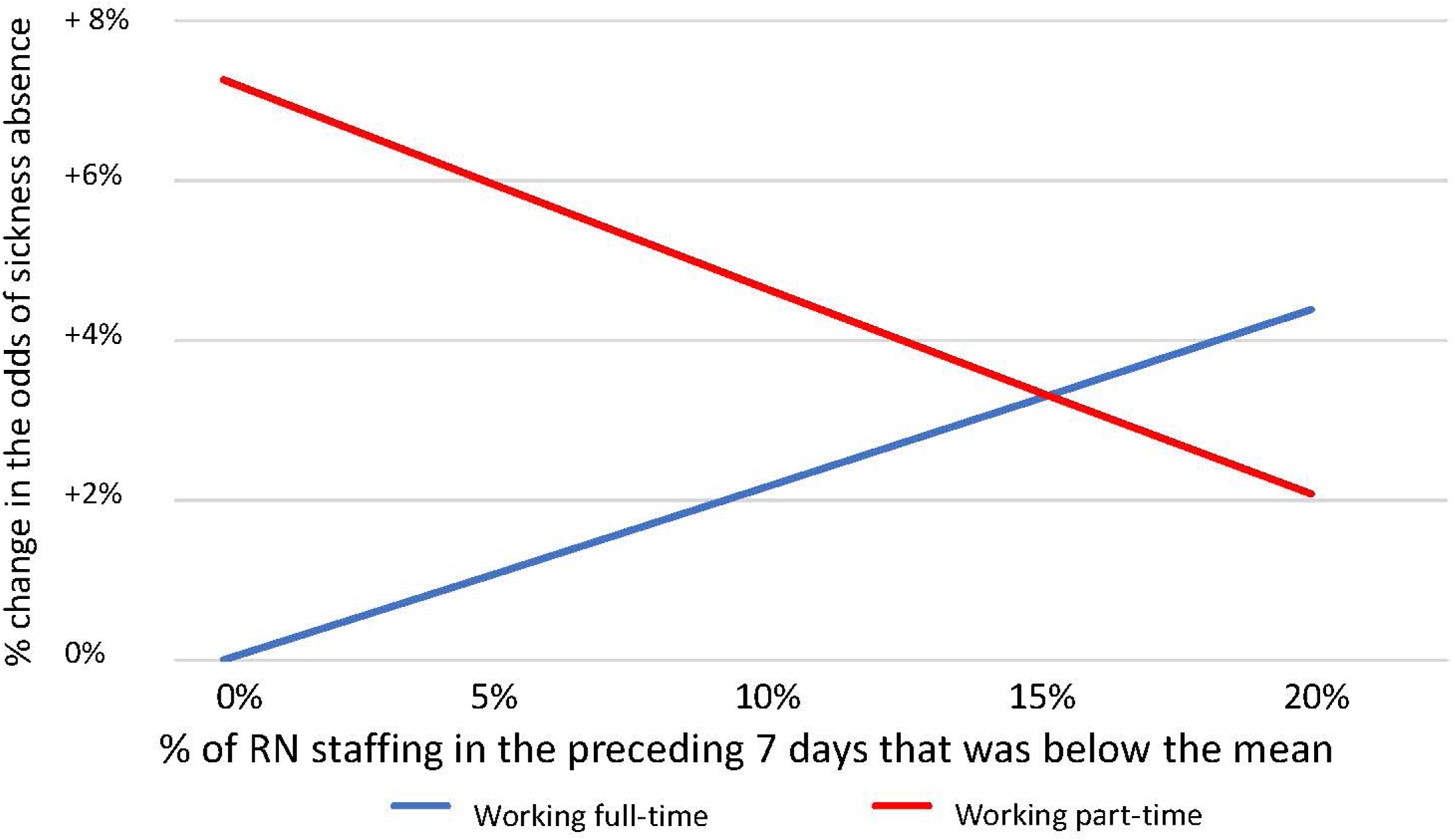
Interaction between RN understaffing and part-time work. Change in the odds of sickness absence associated with variation in staffing levels, relative to the mean by part-time/full-time status

The odds of sickness absence were increased when RNs were exposed to higher proportions of RN understaffing and working full-time, while working part time reverses the association between RN understaffing and sickness absence. All other interactions we tested for were not statistically significant.

For NSs, in the parsimonious reduced model, only exposure to RN understaffing (for every 10 percentage points increase in RN understaffing: OR = 1.030; 95% CI = 1.014 – 1.047) and bank hours (for every 10 percentage points increase in exposure to bank hours on the word: OR = 0.957; 95% CI = 0.938 – 0.976) in the past week were predictors of sickness absence.

## Discussion

This was the first study to analyse the association between nurse staffing configurations and nurses’ sickness absence using objective data extracted from hospital systems. With data collected over 5 years in 4 hospitals, we uncovered statistically significant associations between a number of staffing variables and sickness absence. Our studies revealed four key findings: first, we found that RN understaffing in the preceding 7 days was associated with sickness absence for NSs, but for RNs the association was only seen in those working full time. Second, we found being exposed to higher proportions of hours worked by bank nursing staff was associated with lower sickness absence for RNs and NSs. Third, RNs working shifts with a skill-mix richer in RNs in the preceding 7 days were less likely to experience sickness absence. Lastly, RNs working higher proportions of long shifts in the preceding period were more likely to go off sick. These findings are significant because the nursing workforce globally is under increasing pressure with higher proportions of registered nurses experiencing stress-related sickness and leaving their jobs as a consequence and not enough entering the workforce.^14,15^

Our results contribute to the list of known harms of RN understaffing and diluted skill-mixes which have currently mainly focused on patient outcomes.^16,17^

Those working part time (18% of our sample shifts were by nurses classified as working part-time) do not appear to experience the adverse effects of low RN staffing, indeed low staffing appears to reduce their risk of sickness. This may be in part an artefact, because these staff will be working fewer days and so the variable expressed as a proportion relates to fewer days than it does for full-time staff. The finding that part-time work is associated with higher sickness absence is novel, and might stem from the healthy worker effect, whereby workers who are fit and healthy are likely to work more hours.^18^ A form of healthy worker effect may also explain the counter intuitive decrease in sickness when exposed to understaffing for this group, because if healthy they also have more capacity to work increased hours when staffing is challenged.

RN understaffing and diluted skill-mixes might lead to increased pressure, stress, higher levels of burnout, lower job satisfaction within nursing staff^19,20^ because RNs are responsible for a number of complex activities that cannot be delegated to NSs. Deploying enough or more NSs to counter RN understaffing does not appear to be an effective solution to relieve the pressure on the nursing team. Higher nursing staff stress and pressure might act as mediators in the relationship between RN understaffing and sickness absence, creating a negative vicious circle where staff wellbeing declines and sickness absence increases. Increasing the proportion of NSs might lead to capacity issues and more stress and pressure on RNs because they cannot adequately support and supervise NSs, meaning that the assumed benefits from increasing overall nursing numbers are not realised and the RN workforce is negatively affected. This is an important finding at a time when challenges around employing enough RNs are driving the deployment of a more diluted skill mix.^21^

Relatedly, we did not find any evidence of adverse effects from low NS staffing that mirrors the effects from RN staffing - emphasising that low RN staffing levels are the central problem that should be addressed. In the face of low NS staffing, RNs can flex their role to cover gaps whereas NSs are unable to do the same to cover for RN shortages.

Although high use of temporary staff has been associated with adverse outcomes for staff and patients in previous research, ^22^ ^23^ we did not see clear evidence of any such effect. Although working with a high proportion of agency staff in the previous week increased the risk of sickness absence this was not statistically significant. It is possible that high use of these temporary staff results from successful attempts to manage understaffing, although we did not observe an interaction between the two variables. High use of temporary bank staff was associated with reduced sickness absence. Again, interaction with understaffing seems plausible but was not observed. In view of previous findings this warrants further study.

When registered nurses worked high proportions of long shifts of 12+ hours they were more likely to experience sickness absence. This has been observed previously.^5,6^ Our study strengthens previous findings thanks to its larger and more diverse sample. It also corroborates the hypothetical mechanism whereby long shifts lead to higher cumulative fatigue,^24^ meaning that the resulting extra days off do not constitute an adequate recovery mechanism, and staff experience sickness absence as a result. In addition, a prospective long shift might raise the threshold for an individual worker to decide to attend work if they are not feeling well enough, while a shorter shift might appear more manageable even if feeling unwell.

## Limitations

While our study is longitudinal and we consider a number of working conditions staff are exposed to before experiencing sickness absence, the negative findings around RN understaffing, diluted skill-mixes and long shifts might be reflective of poor working environments, e.g., a good ward environment has simultaneously better RN staffing levels, fewer long shifts and sickness levels, but our data do not allow us to disentangle this. Nonetheless, our findings are similar to those of other studies where low staffing, long shifts and diluted skill- mixes are consistently associated with negative outcomes for patients and staff.

While sickness absence is an objective indicator of staff behaviour, it is multifactorial in nature and we did not have access to the main reason of sickness absence, or any other demographics that could influence an employee’s likelihood of experiencing sickness absence. Nonetheless, we were able to cluster sickness absence episodes within individuals, meaning that personal characteristics were partially controlled for.

## Conclusions

When a clustering of adverse staffing configurations – low RN staffing levels, skill-mixes that are poorer in RNs, working high proportions of long shifts – occurs, the consequences for nursing staff are severe, and their odds of experiencing sickness absence increase. In a climate where workforce wellbeing, efficiency and productivity are high on health systems’ agendas, those in charge of planning workforce at strategic and local levels should invest in employing more registered nurses in hospital settings. Increasing registered nurse staffing levels has been associated with improved outcomes for patients and reduced costs,^25^ and our study adds to this body of evidence by shining a light on the additional impact on nurse health and wellbeing. Given the considerable costs associated with sickness absence, investing in registered nurses has the potential to reduce the pressure on an already exhausted workforce and health system.

## Data Availability

No data can be shared. Unfortunately, due to the sensitive nature of the data and data sharing agreements with the providers, we are unable to freely share the source data, but we guarantee its authenticity and the rigour of methods used in the analysis.

## Author Contributions

CDO and PG conceptualised the study. CDO, PG and JJ contributed to funding acquisition. PM and CS contributed to data curation. CDO, PM, CS, JJ, PG contributed to methodology. CDO, PM, CS, JJ, PG contributed to analysis and interpretation of data. CDO drafted the paper.

All authors have read and approved the final submitted version. CDO acts as guarantor. The corresponding author attests that all listed authors meet authorship criteria and that no others meeting the criteria have been omitted.

The authors would like to acknowledge the contributions of the following co-applicants to the main study (NIHR Project reference: NIHR128056) which gave rise to this sub cohort analysis, in particular for their conceptualisation of and funding acquisition for that study:

Jane Ball^1^

David Culliford^1^

Francesca Lambert^1^

Bruna Rubbo^1,2^

^1^ University of Southampton

^2^ University of Bristol

## Competing interests

All authors have completed the ICMJE uniform disclosure form at www.icmje.org/disclosure-of-interest/ and declare: support from the National Institute for Health and Care Research (NIHR) Health Services and Delivery Research programme, and NIHR Applied Research in Collaboration (Wessex) for the work reported; no financial relationships with any organisations that might have an interest in the submitted work in the previous three years; no other relationships or activities that could appear to have influenced the submitted work.

## Funding statement

The project was funded by the National Institute for Health and Care Research (NIHR) Health Services and Delivery Research Programme (award No. NIHR128056) and the NIHR Applied Research Collaboration (Wessex).

The funder agreed to the protocol but had no part in the design and conduct of the study; collection, management, analysis, and interpretation of the data; preparation, review, or approval of the manuscript; and decision to submit the manuscript for publication.

## Disclaimer

This paper presents independent research commissioned by the NIHR. The views and opinions expressed by authors in this publication are those of the authors and do not necessarily reflect those of the National Health Service, the NIHR Coordinating Centre, the Health and Social Care Delivery Research Programme or the Department of Health and Social Care

## Data Access, Responsibility and Analysis

CDO, PG and PM had full access to all the data in the study and take responsibility for the integrity of the data and the accuracy of the data analysis.

That study was pre-registered at ClinicalTrials.gov (Identifier NCT04374812). The study was funded by the National Institute of Health Research and the full study protocol is available at https://www.fundingawards.nihr.ac.uk/award/NIHR128056.

## Notes

### Author Declarations

This study received ethical approval by the Health Research Authority (IRAS 273185) and the University of Southampton ethics committee (ERGO 52957).

